# A Phenome-Wide Association Study of genes associated with COVID-19 severity reveals shared genetics with complex diseases in the Million Veteran Program

**DOI:** 10.1101/2021.05.18.21257396

**Authors:** Anurag Verma, Noah Tsao, Lauren Thomann, Yuk-Lam Ho, Sudha Iyengar, Shiuh-Wen Luoh, Rotonya Carr, Dana Crawford, Jimmy T. Efird, Jennifer Huffman, Adriana Hung, Kerry Ivey, Michael Levin, Julie Lynch, Pradeep Natarajan, Saiju Pyarajan, Alexander Bick, Lauren Costa, Giulio Genovese, Richard Hauger, Ravi Madduri, Gita Pathak, Renato Polimanti, Benjamin Voight, Marijana Vujkovic, Maryam Zekavat, Hongyu Zhao, Marylyn D Ritchie, VA Million Veteran Program COVID-19 Science Initiative, Kyong-Mi Chang, Kelly Cho, Juan P. Casas, Philip S. Tsao, J. Michael Gaziano, Christopher O’Donnell, Scott Damrauer, Katherine Liao

## Abstract

The study aims to determine the shared genetic architecture between COVID-19 severity with existing medical conditions using electronic health record (EHR) data. We conducted a Phenome-Wide Association Study (PheWAS) of genetic variants associated with critical illness (n=35) or hospitalization (n=42) due to severe COVID-19 using genome-wide association summary from the Host Genetics Initiative. PheWAS analysis was performed using genotype-phenotype data from the Veterans Affairs Million Veteran Program (MVP). Phenotypes were defined by International Classification of Diseases (ICD) codes mapped to clinically relevant groups using published PheWAS methods. Among 658,582 Veterans, variants associated with severe COVID-19 were tested for association across 1,559 phenotypes. Variants at the *ABO* locus (rs495828, rs505922) associated with the largest number of phenotypes (n_rs495828_= 53 and n_rs505922_=59); strongest association with venous embolism, odds ratio (OR_rs495828_ 1.33 (p=1.32 × 10^−199^), and thrombosis OR_rs505922_ 1.33, p=2.2 ×10^−265^. Among 67 respiratory conditions tested, 11 had significant associations including *MUC5B* locus (rs35705950) with increased risk of idiopathic fibrosing alveolitis OR 2.83, p=4.12 × 10^−191^; *CRHR1 (*rs61667602) associated with reduced risk of pulmonary fibrosis, OR 0.84, p=2.26× 10^−12^. The *TYK2* locus (rs11085727) associated with reduced risk for autoimmune conditions, e.g., psoriasis OR 0.88, p=6.48 ×10^−23^, lupus OR 0.84, p=3.97 × 10^−06^. PheWAS stratified by genetic ancestry demonstrated differences in genotype-phenotype associations across ancestry. *LMNA* (rs581342) associated with neutropenia OR 1.29 p=4.1 × 10^−13^ among Veterans of African ancestry but not European. Overall, we observed **a** shared genetic architecture between COVID-19 severity and conditions related to underlying risk factors for severe and poor COVID-19 outcomes. Differing associations between genotype-phenotype across ancestries may inform heterogenous outcomes observed with COVID-19. Divergent associations between risk for severe COVID-19 with autoimmune inflammatory conditions both respiratory and non-respiratory highlights the shared pathways and fine balance of immune host response and autoimmunity and caution required when considering treatment targets.

## Introduction

Coronavirus disease 2019 (COVID-19) first identified in December of 2019^1^, became a global pandemic by March 2020. As of September 2021, COVID-19, transmitted by the SARS-CoV-2 virus, has resulted in the loss of over 4.6 million lives worldwide.^2^ Identifying host genetic variants associated with severe clinical manifestations from COVID-19, can identify key pathways important in the pathogenesis of this condition. International efforts such as the COVID-19 Host Genetics Initiative (HGI)^3^ have meta-analyzed genome-wide association study (GWAS) summary statistics at regular intervals to identify novel genetic associations with COVID-19 severity. Thus far, ten independent variants associated with COVID-19 severity at genome-wide significance have been identified, most notably at the *ABO* locus.^4^ These GWASs have also identified variations in genes involving inflammatory cytokines and interferon signaling pathways such as *IFNAR2, TYK2*, and *DPP9*.^4^

The unprecedented availability of genome-wide data for COVID-19 provides an opportunity to study clinical conditions that share genetic risk factors for COVID-19 severity. Examining known conditions, each with a body of knowledge regarding important pathways and targets, may in turn improve our understanding of pathways relevant for COVID-19 severity and inform the development of novel treatments against this pathogen. The Phenome-Wide Association Study (PheWAS) is an approach for simultaneously testing genetic variants’ association with a wide spectrum of conditions and phenotypes.^5^ The Veteran’s Affairs (VA) Million Veterans Program (MVP) has generated genotypic data on over 650,000 participants linked with electronic health record (EHR) data containing rich phenotypic data, enables large-scale PheWAS. Moreover, MVP has the highest racial and ethnic diversity of the major biobanks worldwide affording an opportunity to compare whether associations are similar across genetic ancestries.^6^

The objective of this study was to use existing clinical EHR data to identify conditions that share genetic variants with COVID-19 severity using the disease-agnostic PheWAS approach. Since COVID-19 is a new condition, identifying existing conditions which share genetic susceptibility may allow us to leverage existing knowledge from these known conditions to provide context regarding important pathways for COVID-19 severity, as well as how pathways may differ across subpopulations.

## Methods

### Data sources

The VA MVP is a national cohort launched in 2011 designed to study the contributions of genetics, lifestyle, and military exposures to health and disease among US Veterans.^6^ Blood biospecimens were collected for DNA isolation and genotyping, and the biorepository was linked with the VA EHR, which includes diagnosis codes (International Classification of Diseases ninth revision [ICD-9] and tenth revision [ICD-10]) for all Veterans followed in the healthcare system up to September 2019. The single nucleotide polymorphism (SNP) data in the MVP cohort was generated using a custom Thermo Fisher Axiom genotyping platform called MVP 1.0. The quality control steps and genotyping imputation using 1000 Genomes cosmopolitan reference panel on the MVP cohort has been described previously.^7^ All individuals in the study provided written informed consent as part of the MVP. This study was approved through the Veterans Affairs central institutional review board as part of the MVP.

#### Genetic variant selection

An overview of the analytic workflow is outlined in Fig 1. Variants were derived from the COVID-19 HGI GWAS meta-analysis release v6^3^. In this study, we analyzed the following HGI GWAS summary statistics: 1) hospitalized and critically ill COVID-19 vs. population controls denoted as “A2” in HGI, and referred to as “critical COVID” in this study, and 2) hospitalized because of COVID-19 vs. population controls, denoted as “B2” in HGI, referred to as “hospitalized COVID” in this study^3^. For each GWAS, variants with a Benjamini-Hochberg false discovery rate (FDR) corrected p-value < 0.01 were selected as candidate lead SNPs (3,502 associated with critical COVID, and 4,336 associated with hospitalized COVID). Variants with r^2^ <0.1 were clustered within a 250 kb region according to 1000 Genomes phase 3 transethnic reference panel^8^, resulting in 45 independent variants associated with critical COVID and 42 variants associated with hospitalized COVID summary statistics. The lead variants from each set of GWAS summary statistics are available in eTable 1.

**Table 1.** Patient characteristics of Million Veteran Program participants

| Characteristics | Million Veteran Program |
| --- | --- |
|  | Number (%) |
| Total Patients | 658,582 |
| Male | 592,516 (90) |
| Genetic Ancestry |  |
| European | 464,961 (70) |
| African | 123,120 (19) |
| Hispanic | 52,183 (8) |
| Asian | 83,29 (1) |
| Other | 99,89 (2) |
| Comorbidities |  |
| Obesity (phecode = 278) | 283,197 (43) |
| Hypertension (phecode = 401.1) | 451,998 (69) |
| Type 2 Diabetes (phecode = 250.2) | 227,575 (34) |
| Coronary Artery Disease (phecode = 411.4) | 152,136 (23) |
| Chronic Kidney Disease (phecode = 585.2) | 100,46 (15) |

**Fig 1.**
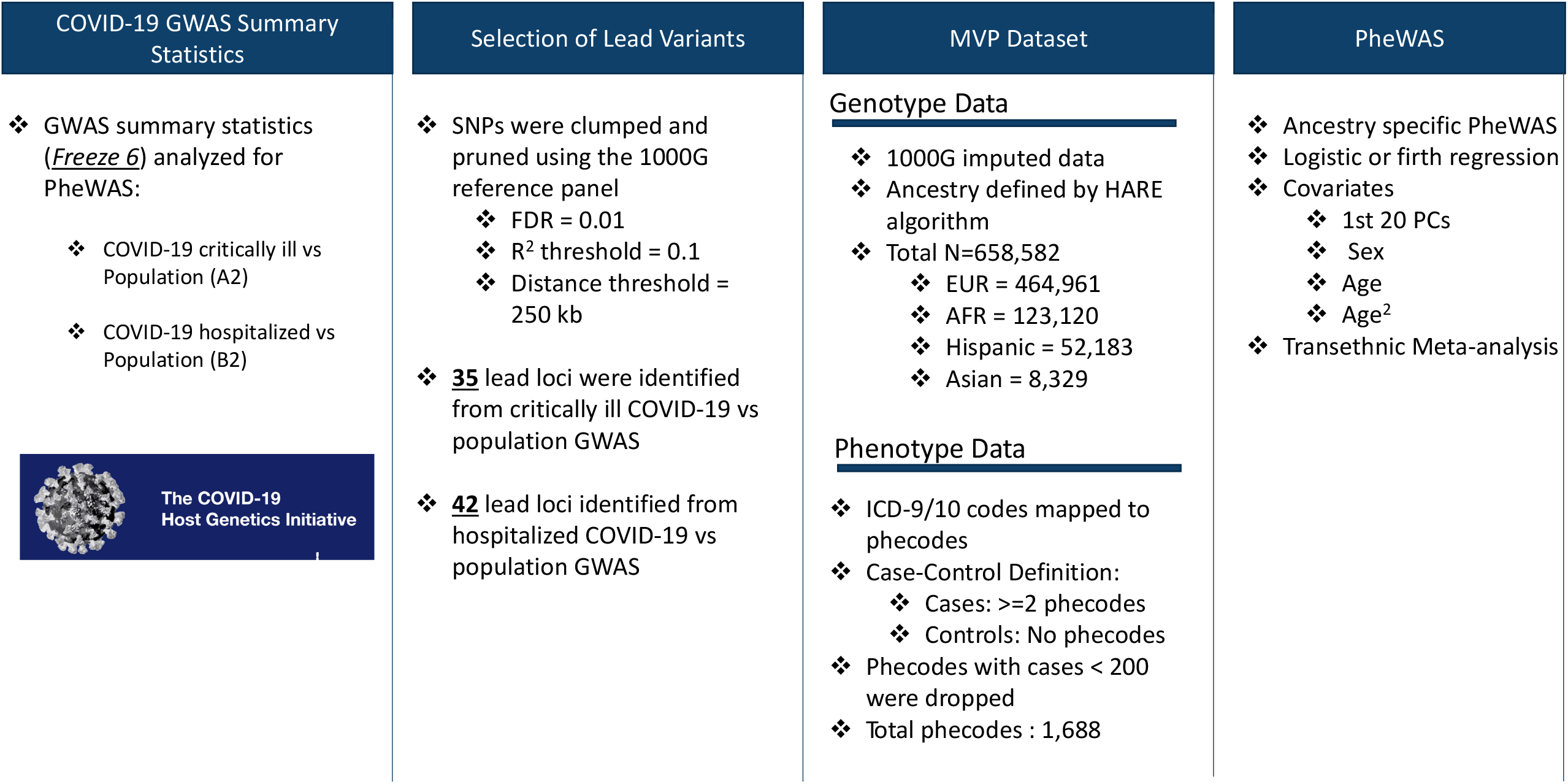
Overview of variant selection and PheWAS analysis design.

#### Outcomes

For both MVP, clinical data prior to the onset of the COVID-19 pandemic were used to reduce potential confounding bias from SARS-CoV-2 infection on existing conditions. Phenotypes were defined by phecodes from prior studies^5,9^. Each phecode represents ICD codes grouped into clinically relevant phenotypes for clinical studies. For example, the phecode “deep venous thrombosis” includes “venous embolism of deep vessels of the distal lower extremities,” and “deep venous thrombosis of the proximal lower extremity,” both of which have distinct ICD codes. Using this approach, all ICD codes for all Veterans in MVP were extracted and each assigned a phenotype defined by a phecode. ICD-9 and ICD-10 codes were mapped to 1876 phecodes, as previously described.^5,9^

For each phecode, participants with ≥2 phecode-mapped ICD-9 or ICD-10 codes were defined as cases, whereas those with no instance of a phecode-mapped ICD-9 or ICD-10 code were defined as controls. Based on our previous simulation studies of ICD EHR data, populations where the phecode comprises < 200 cases were more likely to result in spurious results^10^, and we thus applied this threshold in each ancestry group. In total, we analyzed 1,617 (EUR), 1304 (AFR), 993 (HIS), 294 (ASN) phecodes from the MVP cohort.

#### Phenome-wide association studies

The primary PheWAS analysis used SNPs identified from the HGI GWAS of critical and hospitalized COVID, and tested association of these SNPs with phenotypes extracted from the EHR using data prior to the COVID-19 pandemic. Logistic regression using PLINK2 to examine the SNP association with phecodes and firth regression was applied when logistic regression model failed to converge. Regression models were adjusted for sex, age (at enrollment), age squared, and the first 20 principal components. Genetic ancestry was determined using the HARE method for four major groups: African (AFR), Asian (ASN), Hispanic (HIS), and European (EUR) ancestry^11^. Ancestry-specific PheWAS was first performed in these four groups, and summary data were meta-analyzed using an inverse-variance weighted fixed-effects model implemented in the PheWAS R package^9^. We assessed heterogeneity using I^2^ and excluded any results with excess heterogeneity (I^2^ > 40%).

To address multiple testing, an association between SNP and phecode with FDR p < 0.01 was considered significant. Thus, the threshold for significance was p < 6.07 × 10^−05^ for critical COVID lead variants, and p < 4.13 × 10^−05^ for hospitalized COVID lead variants. In the main manuscript we highlight PheWAS significant associations using FDR < 0.01 and an effect size associated with increased or reduced risk for a condition by 10%, with complete PheWAS results provided in S2 Table and S3 Table.

## Results

We studied 658,582 MVP participants, with mean age 68 years (SD), 90% male, with 30% participants from non-European ancestry (Table 1). The PheWAS was performed on 35 genetic variants associated with critical COVID-19, and 42 genetic variants (S1 Table) associated with hospitalized COVID, across 1,559 phenotypes.

From the trans-ethnic meta-analysis, we identified 151 phenotypes significantly associated with critical COVID GWAS-identified variants, and 156 associations with hospitalized COVID GWAS-identified lead variants (FDR, p<0.01). Among these lead variants with significant PheWAS associations, 10 SNPs were associated with reduced risk of critical and hospitalized COVID-19 in HGI. Six variants were common to both severe and hospitalized COVID and had significant PheWAS associations, namely, variations nearest to the genes *ABO* (rs495828 and rs505922), *DPP9* (rs2277732), *MUC5B* (rs35705950), TYK2 (rs11085727), and *CCHCR1* (rs9501257) (S2 Table and S3 Table).

### Association of ABO loci with known risk factors and outcomes related to COVID-19 severity

In the transethnic meta-analysis, the phenotypes with the strongest association with variants near *ABO* locus (rs495828 and rs505922) was “hypercoagulable state” (OR_critical_PheWAS_ = 1.48 [1.42 - 1.54], *P*_critical_PheWAS_ = 1.84 × 10^−40^; OR_hospitalized_PheWAS_ = 1.51 [1.46 - 1.56], *P*_hospitalized_PheWAS_ = 2.11 × 10^−55^, Fig 2). The ABO loci had the largest number of significant PheWAS association findings, accounting for 35% (53/151) of significant phenotype associations in the critical COVID PheWAS, and 37% (59/156) in the hospitalized COVID PheWAS. The phenotypes with the most significant associations and largest effect size were related to hypercoagulable states and coagulopathies. As expected, conditions not related to coagulopathy associated with the *ABO* locus, included type 2 diabetes and ischemic heart disease, have been reported as risk factors for or are complications associated with COVID-19 severity and mortality (Fig 2, S2 Table and S3 Table).

**Fig 2.**
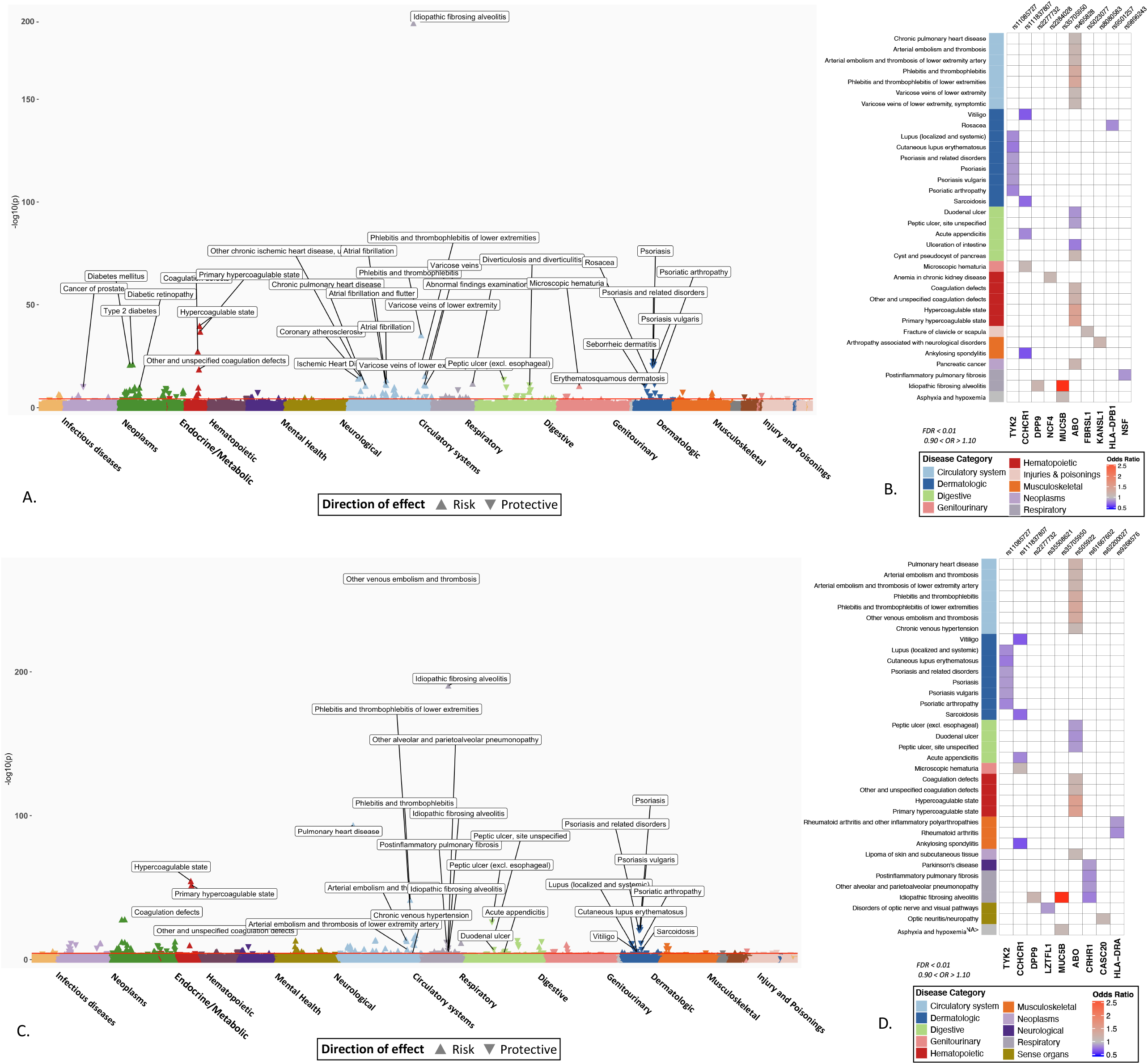
PheWAS results of candidate SNPs from GWAS of critically ill and hospitalized COVID-19. Significant associations between 48 SNPs from critical ill COVID GWAS (A) and 39 SNPs from hospitalized COVID (C) and EHR derived phenotypes in the Million Veteran Program. The phenotypes are represented on the x-axis and ordered by broader disease categories. The red line denotes the significance threshold using false discovery rate of 1% using the Benjamini-Hochberg procedure. The description of phenotypes is highlighted for the associations with FDR < 0.1 and odds ratio < 0.90 or odds ratio > 1.10. B) and D) A heatmap plot of SNPs with at least one significant association (FDR < 0.1). The direction of effect disease risk is represented by odds ratio. A red color indicates increased risk and blue color indicated reduced risk. The results with odds ratio < 0.90 or odds ratio > 1.10 are shown.

### Associations between variants associated with COVID-19 severity and respiratory conditions and infections

Among 68 respiratory conditions, only 11 diseases had significant associations (FDR < 0.01) shared with genetic variants associated with severe COVID-19. The most significant association was observed between rs35705950 (*MUC5B*) and idiopathic fibrosing alveolitis (OR = 2.83 [2.76 – 2.90]; *P* = 4.12 × 10^−191^), also known as idiopathic pulmonary fibrosis (IPF). Similarly, rs2277732 near *DPP9* was associated with IPF (OR = 1.16 [1.09 – 1.22]; *P* = 5.84 × 10^−06^), both association between *MUC5B, DPP9* variants and IPF has been reported in previous studies.^12^ However, the association of genetic variants with other respiratory conditions may represent novel findings: the association of intronic variant rs61667602 in *CRHR1* with reduced risk of post inflammatory pulmonary fibrosis (OR = 0.84 [0.80 – 0.89]; *P* = 2.26× 10^−12^), “alveolar and parietoalveolar pneumonopathy” (OR = 0.80 [0.72 – 0.88]; *P* = 1.58 × 10^−08^) and IPF (OR = 0.87 [0.82 - 0.92], *P* = 7.5 × 10^−07^). We did not detect associations between any of the variants and other respiratory conditions which are known risk factors for COVID-19 such as COPD, cystic fibrosis, pulmonary hypertension. (S2 Table, S3 Table).

### Associations between variants associated with COVID-19 severity and reduced risk for certain phenotypes

The rs11085727-T allele of *TYK2*, a lead variant from the both critically ill and hospitalized COVID GWAS was associated with a reduced risk for psoriasis (OR = 0.88 [0.86-0.91], *P* = 6.48 × 10^−23^), psoriatic arthropathy (OR = 0.82 [0.76 - 0.87], *P* = 6.97 × 10^−12^), and lupus (OR = 0.84 [0.76 - 0.91], *P* = 63.97 × 10^−06^). This *TYK2* signal has been previously reported to be associated with reduced risk of psoriasis, psoriatic arthropathy, type 1 diabetes, systemic lupus erythematosus and RA as well as other autoimmune inflammatory conditions^13,14^ (Table 2).

**Table 2.** Phenotypes sharing association with variants also associated with severe COVID-19 infection, with reduced odds of disease listed in order of p-value*.

| Phenotype | OR (95% CI) | p-value | Gene | SNP | COVID-severity |
| --- | --- | --- | --- | --- | --- |
| Psoriasis | 0.89 [0.86-0.91] | 6.48E-23 | *TYK2* | rs11085727 | Both |
| Rosacea | 0.84 [0.8-0.89] | 7.54E-16 | *HLA-DPB1* | rs9501257 | Critical |
| Psoriatic arthropathy | 0.82 [0.77-0.88] | 6.97E-12 | *TYK2* | rs11085727 | Both |
| Post-inflammatory pulmonary fibrosis | 0.87 [0.83-0.92] | 4.54E-09 | *NSF* | rs9896243 | Critical |
| Vitiligo | 0.69 [0.56-0.82] | 3.03E-08 | *CCHCR1* | rs111837807 | Both |
| Sarcoidosis | 0.74 [0.62-0.85] | 1.80E-07 | *CCHCR1* | rs111837807 | Both |
| Lupus (localized and systemic) | 0.84 [0.77-0.91] | 3.97E-06 | *TYK2* | rs11085727 | Both |
| Cutaneous lupus erythematosus | 0.79 [0.68-0.89] | 6.21E-06 | *TYK2* | rs11085727 | Both |
| Post-inflammatory pulmonary fibrosis | 0.85 [0.8-0.9] | 2.26E-12 | *CRHR1* | rs61667602 | Hospitalized |
| Rheumatoid arthritis | 0.84 [0.79-0.9] | 4.20E-10 | *HLA-DRA* | rs9268576 | Hospitalized |
| Idiopathic fibrosing alveolitis | 0.81 [0.73-0.88] | 1.58E-08 | *CRHR1* | rs61667602 | Hospitalized |
| Rheumatoid arthritis and other inflammatory polyarthropathies | 0.88 [0.84-0.93] | 6.34E-08 | *HLA-DRA* | rs9268576 | Hospitalized |
| Other alveolar and parietoalveolar pneumonopathy | 0.88 [0.83-0.93] | 7.50E-07 | *CRHR1* | rs61667602 | Hospitalized |

\*OR<0.9 and P<10<sup>-5</sup> shown in table, full results in supplementary; if multiple related conditions, e.g. psoriasis, psoriasis vulgaris, psoriasis and related disorders, description with lowest p-value selected shown in table.

### Ancestry specific PheWAS provide insights into disease risks across ancestries

The PheWAS analyses performed across four major genetic ancestry group in MVP observed similar findings as the overall meta-analysis with few associations unique to each ancestry. (Fig 3, S8 Table). SNP rs581342 (*LMNA*), associated with severe COVID-19, was a highly prevalent variant among subjects with AFR ancestry (MAF=0.53) and was associated with neutropenia (OR_AFR_ = 0.82 [0.76 - 0.87], *P*_*AFR*_ = 4.09 × 10^−13^); this association was not observed in larger population of EUR descent (S8 Table). Following up on this finding, we extracted data on laboratory values and observed a strong association between *LMNA* with lower white blood cell count (beta = -0.34 [-0.35, -0.32], *P*_AFR_= 1 × 10^−300^) and lower median neutrophil fraction (beta = -1.84 [-1.94, -1.75], *P*_AFR_ = 1 × 10^−300^) compared to those without this variant. This association in laboratory values was again more significant with a stronger effect size among subjects with AFR ancestry in comparison to EUR (P=0.005). Among AFR individuals, each allele was associated with a 1.84% lower neutrophil fraction, where among EUR individuals, each allele was associated with only a 0.04% reduction (S9 Table).

**Fig 3.**
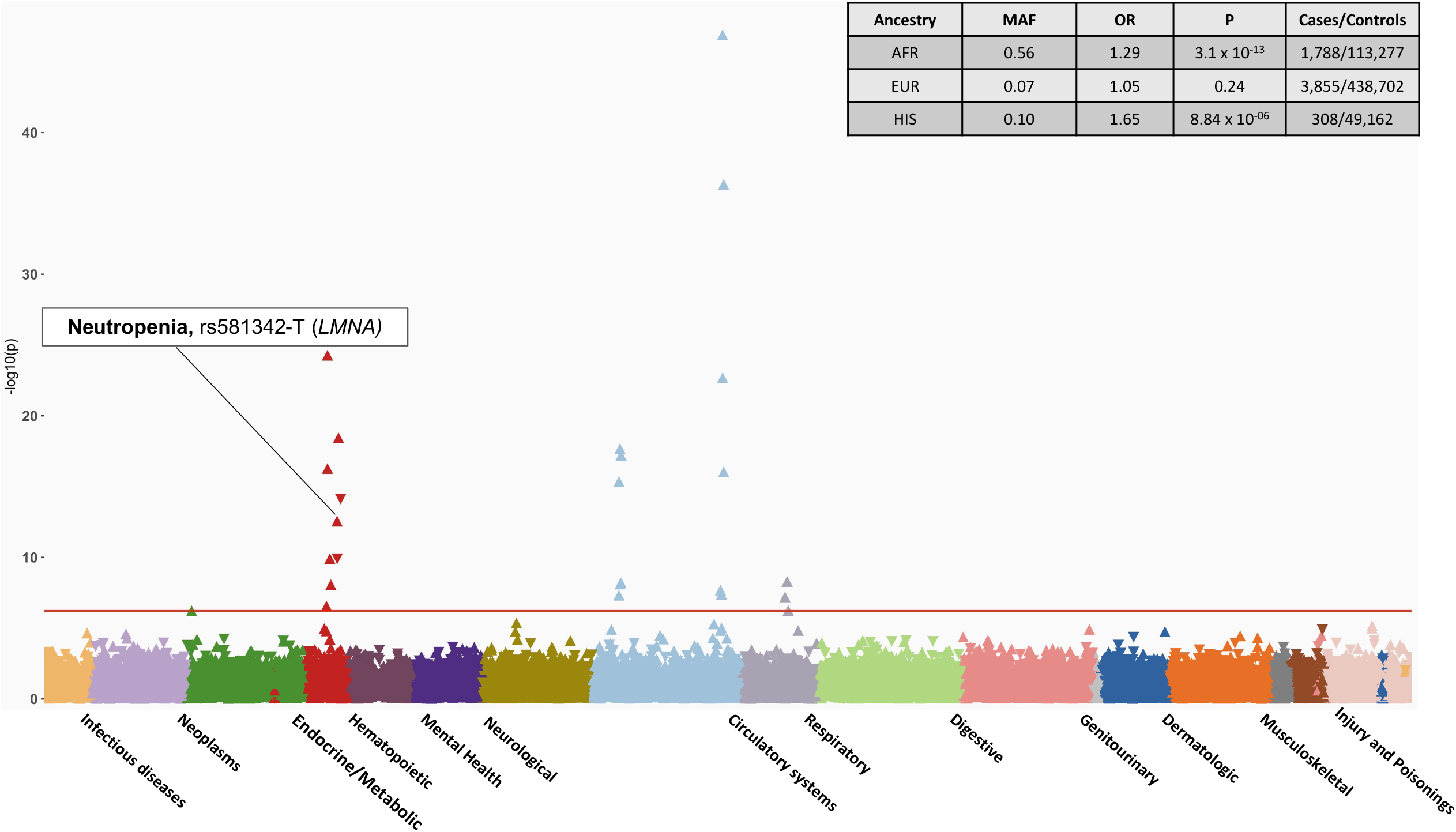
PheWAS results of candidate SNPs from GWAS of Hospitalized COVID-19 in AFR ancestry individuals. The plot highlights the association between rs581342 SNP and Neutropenia, which was only observed in the AFR ancestry. The phenotypes are represented on the x-axis and ordered by broader disease categories. The red line denotes the significance threshold using false discovery rate of 1% using the Benjamini-Hochberg procedure. The table on the top right of the plot shows the association results between rs581342 and neutropenia in other ancestries. The association was not tested among participants of ASN ancestry due to low case numbers.

Similarly, associations between rs9268576 (*HL-DRA*) and thyrotoxicosis was only observed in AFR ancestry participants. The EUR ancestry specific PheWAS identified 39 significant associations which were not observed in other ancestry groups. One such association was between MUC5B variant and phecode for “dependence on respirator [Ventilator] or supplemental oxygen” (OR_EUR_ = 1.16 [1.11 – 1.12], *P*_*EUR*_ = 1.72× 10^−10^) among EUR ancestry participants was not significant in other ancestry population (S8 Table). It is important to note that the conditions with significant association among EUR participants had similar prevalence among other ancestries. However, since there were overall fewer subjects in non-EUR ancestry groups, this likely resulted in lower statistical power to detect associations. All ancestry specific PheWAS results are available in supplementary tables (S4 Table, S5 Table, S6 Table, S7 Table).

### Association with variation at sex chromosome

In the hospitalized COVID-19 GWAS, we identified rs4830964 as the only lead variant on chromosome X. The SNP is located near *ACE2* and was associated “non-healing surgical wound” (OR = 0.92 [0.89 – 0.96], *P* = 2.23× 10^−05^). Notably, the SNP had nominal association (p<0.05) with type 2 diabetes and diabetes related complications that are previously reported association with variation in *ACE2* (S3 Table). We did not observe any association with this variant in the ancestry specific PheWAS analysis.

## Discussion

In this large-scale PheWAS, we identified the shared genetic architecture between variants associated with severe COVID-19 and other complex conditions using data from MVP, one of the largest and most diverse biobanks in the world. Broadly, these risk alleles identified conditions associated with risk factors for severe COVID-19 manifestations such as T2D, ischemic heart disease across all ancestries. Notably, the strongest associations with the highest effect size were related to coagulopathies, specifically, hypercoagulable state including deep venous thrombosis and other thrombotic complications, also shared variants associated with severe COVID-19. In contrast, among respiratory conditions, only idiopathic pulmonary fibrosis and chronic alveolar lung disease shared genetic risk factors, with the notable absence of an association with COPD, pulmonary hypertension, and other respiratory infections. When comparing findings from the two largest ancestry groups in MVP, AFR and EUR, we observed that a risk allele associated with severe COVID-19 that shares an association with neutropenia on among Veterans of AFR ancestry. Finally, we observed that variants associated with severe COVID-19 had an opposite association, or reduced odds with autoimmune inflammatory conditions, such as psoriasis, psoriatic arthritis, RA, and inflammatory lung conditions.

A classic GWAS tests the association between millions of genetic variants with the presence or absence of one phenotype, e.g., GWAS of deep venous thrombosis. In the COVID-19 HGI GWAS, the “phenotype” was patients hospitalized for or critically ill from COVID-19. Clinically, this population includes a mixture of patients with a complex list of medical conditions at high risk for severe COVID complications and those who had actual complications from COVID-19. Thus, we would anticipate that many of the significant phenotypes would be associated with risk factors such as obesity and deep venous thrombosis. The clinical data used in this study pre-dates the emergence of COVID-19 to reduce potential confounding bias that can occur in a population infected with SARS-CoV-2, e.g., interaction between COVID-19 and type 2 diabetes. Additionally, our findings suggest that the PheWAS approach can be a useful tool to identify clinical factors related to emerging infectious diseases regarding severity or complications when genomic data are available.

The PheWAS results of SNPs in the *ABO* locus served as a positive control for this study. Genetic variations in *ABO* are an established risk factor for COVID-19 severity. Patients with blood group A have a higher risk of requiring mechanical ventilation and extended ICU stay compared with patients with blood group O.^15^ These same variations at *ABO* had known associations with a spectrum of blood coagulation disorders identified in studies pre-dating COVID-19.^16–18^ The PheWAS of ABO variants identified associations with increased risk of deep vein thrombosis, pulmonary embolism, and other circulatory disorders, in line with prior studies, and recent studies among patients hospitalized with COVID-19.^19–23^

Among the respiratory conditions, only idiopathic pulmonary fibrosis (IPF) and chronic alveoli lung disease had associations with the variants near genes *MUC5B, CRHR1*, and *NSF*. Located in the enhancer region of the *MUC5B*, rs35705950, is a known risk factor for IPF, and a high mortality rate was observed among the COVID-19 patients with pre-existing IPF.^24^ However, the variant is associated with a reduced risk of severe COVID-19 (OR=0.89), revealing the risk allele’s opposing effect for infection and pulmonary fibrosis. In a separate study of MVP participants tested for COVID-19, we identified a significant mediating effect of the *MUC5B* variant in reducing risk for pneumonia due to COVID-19^25^. An intronic variation in *CRHR1* (rs61667602-T) had reduced risk for severe COVID-19 (OR= 0.91) as well as respiratory conditions such as IPF. *CRHR1* gene is a receptor that binds to the corticotropin-releasing hormone has a key role in immune, behavioral, autonomic, and neuroendocrine responses to stress. Depression and anxiety are the known conditions associated with variations in *CRHR1*, but variations in this gene have also shown associations enhanced improvement in pulmonary function in asthma patients taking inhaled corticosteroid^26^. This finding may inform results from the RECOVERY clinical trial of patients hospitalized with COVID-19 where a survival benefit was observed for dexamethasone use among those receiving respiratory support^27^.

Several conditions shared genetic variants associated with severe COVID-19, however, the association was for reduced odds for these conditions. All except one, rosacea, have a known autoimmune etiology. The existing literature can help explain the dual association between reduced risk of autoimmune conditions such as psoriasis and RA and increased risk of severe COVID-19 via *TYK2. TYK2*, a member of the Janus Kinase (*JAK*) family of genes, plays a key role in cytokine signal transduction and the inflammatory response, particularly via IL-12, IL-23, and is also important for IL-6 and IL-10 signaling (Fig 3).^28^ *TYK2* serves a central role in type 1 interferon signaling, part of the innate immune response blocking the spread of a virus from infected to uninfected cells. Partial loss of *TYK2* function is associated with reduced risk for several autoimmune disorders such as RA and psoriatic disease, conditions treated with immunosuppressive therapy.^13,29–32^ Humans with complete *TYK2* loss of function have clinically significant immunodeficiency with increased susceptibility to mycobacterial and viral infections.^28,33^ In line with the *TYK2* findings is enhanced steroid responsiveness among patients with asthma carrying the *CRHR1* variant^34^. Here again, a variant associated with severe COVID-19 is associated with a non-COVID phenotype responsive to immunosuppressive therapy. In summary, reviewing the overall signal of opposing associations of variants with COVID-19 and autoimmune conditions, highlights the known fine balance between host immune response and autoimmunity.

While non-white populations are disproportionately affected by COVID-19, the current genetic studies of severe COVID-19 still predominantly consist of individuals from EUR ancestry. MVP has the most racial and ethnic diversity compared to other major biobanks. The availability of linked EHR data provide the opportunity to provide more in-depth studies of genotype-phenotype associations observed from the PheWAS. The GWAS from the HGI provides the most diverse genomic data of COVID-19 consisting of participants from over 25 countries EUR (33% non-EUR samples), enabling identification of variants more prevalent in non-EUR populations. In the present study, we observed that a variant located in the *LMNA* gene locus was associated with neutropenia in AFR ancestry but not in other ancestry groups, including EUR which would have been well powered to detect an association. Furthermore, examination of actual neutrophil percentages measured as part of routine care demonstrated stronger associations in Veterans of AFR ancestry compared to EUR.

*LMNA* variants are associated with a broad spectrum of cardiomyopathies such as dilated cardiomyopathies, familial atrial fibrillation. However, the association with neutropenia has not been previously reported. Neutropenia refers to an abnormally low number of neutrophils cell in the blood, and predisposes to increased risk of infection. Epidemiology studies have shown that lower neutrophil counts are more common in individuals with African Ancestry^36^ and are hypothesized to be a result of selection and generally considered benign. Whether low neutrophil levels may clinically impact COVID-19 outcomes remains to be seen and should be further studied.

### Limitations

We note several limitations. First, the PheWAS was designed as a broad screen to test for potentially clinically relevant associations between genes and phenotypes, with limited power to detect associations among uncommon conditions, and when further stratified by genetic ancestry. Findings from this study suggest that variants associated with severe COVID-19 are also associated with reduced odds of having an autoimmune inflammatory condition. However, the results cannot provide information on the impact of actual SARS-CoV-2 infection in these individuals after diagnosis of an autoimmune disease

## Conclusions

The PheWAS of genetic variants reported to associate with severe COVID-19 demonstrated shared genetic architecture between COVID-19 severity and known underlying risk factors for both severe COVID-19 and poor COVID-19 outcomes, rather than susceptibility to other viral infections. Overall, the associations observed were generally consistent across genetic ancestries, with the exception of a stronger association with neutropenia among Veterans of African ancestry than European ancestry. Notably, only few respiratory conditions had a shared genetic association with severe COVID-19. Among these, variants associated with a reduced risk for severe COVID-19 had an opposite association, with reduced risk for inflammatory and fibrotic pulmonary conditions. Similarly, other divergent associations were observed between severe COVID-19 and autoimmune inflammatory conditions, shedding light on the concept of the fine balance between immune tolerance and immunodeficiency. This balance will be important when considering therapeutic targets for COVID-19 therapies where pathways may control both inflammation and the viral host response.

## Data Availability

Full summary-level association data from the PheWAS from this study is made available as supplementary files.

## Funding

This research is based on data from the Million Veteran Program, Office of Research and Development, Veterans Health Administration, and was supported by award MVP035. This publication does not represent the views of the Department of Veteran Affairs or the United States Government. R.M.C. is supported by NIH grants R01 AA026302 and P30 DK0503060. K.P.L. is supported by NIH P30 AR072577, and the Harold and Duval Bowen Fund.

## Conflict of Interest

RMC has received research support from Intercept Pharmaceuticals, Inc and Merck & Co. MDR is on the scientific advisory board for Goldfinch Bio and Cipherome. CJO is an employee of Novartis Institute for Biomedical Research. PN reports grant support from Amgen, Apple, AstraZeneca, Boston Scientific, and Novartis, personal fees from Apple, AstraZeneca, Blackstone Life Sciences, Genentech, and Novartis, and spousal employment at Vertex, all unrelated to the present work.

## Acknowledgements

We are grateful to our Veterans for their contributions to MVP. Full acknowledgements for the VA Million Veteran Program COVID-19 Science Initiative can be found in the supplementary methods. We would like to thank the Host Genetic Initiative for making their data publicly available (https://www.covid19hg.org/acknowledgements/).

## Notes

### Author Declarations

This research is based on data from the Million Veteran Program, Office of Research and Development, Veterans Health Administration, and was supported by award MVP035. This publication does not represent the views of the Department of Veteran Affairs or the United States Government.

### Summary of Updates

Fixed minor typos and corrections

